# Nurse-led point-of-care ultrasonography with telemedicine review to improve the impact of antenatal care: a formative qualitative study in Kenya

**DOI:** 10.1101/2023.10.12.23296931

**Authors:** Meghan Bruce Kumar, Caleb Mike Mulongo, Lucia Pincerato, MariaVittoria DeVita, Salima Saidi, Yvonne Gakii, GianFranco Morino, Pratap Kumar

## Abstract

The informal settlements of Nairobi have higher neonatal and infant mortality rates than the average for Nairobi. Universal access to important diagnostics like ultrasonography is poor and inequitable due to the high cost of devices and limited availability of skilled sonographers. Recent advances of mobile ultrasound probes connected to smartphones, with or without artificial intelligence support, have improved access to devices; but skills to perform and interpret scans continue to be limited. The SonoMobile intervention involved training nurse-midwives to conduct point-of-care obstetric ultrasound scans in antenatal care clinics in urban informal settlements. Scan data and images were shared, using telemedicine technology, with remote sonographers, who reviewed scan images and data, and provided reports. This qualitative study of 61 respondents from diverse stakeholder groups describes the acceptability, utility and potential sustainability of nurse-led, point-of-care obstetric ultrasonography with telemedicine review. Perceived value of nurse-led obstetric ultrasonography include improving access and affordability of obstetric ultrasonography services, timely identification and referral of high-risk pregnancies, and improving awareness of appropriate antenatal care among underserved populations. The relative affordability of SonoMobile was described as a critical enabler for a business model targeting low- and middle-income segments of the population, and for increasing quality and equity of antenatal care coverage. Areas highlighted for improvement include strengthening supervision of nurse trainees, broadening the scope of nurse training, and development of clear regulatory guidelines for nurse-led obstetric ultrasonography. The study highlights the complex task shifting required to provide universal access to a life-saving technology in an LMIC health system.

## Introduction

Kenya has high national maternal and neonatal mortality rates (MMR, NMR, respectively) at 362 per 100,000 live births and 22 per 1,000 live births respectively,^1^ with neonatal mortality accounting for 56% of the overall infant mortality rate. The sustainable development goals (SDGs) aim to reduce the MMR and NMR to 70 and 12, respectively.^2^ The NMR in Nairobi (the capital and largest city) is 77% higher than the national average; the rate in the informal settlements of Nairobi is 133% higher than that of Nairobi and the MMR almost twice the national average.^3^ Although living in or near Nairobi increases geographic proximity to health facilities, it also brings challenges related to high population density and poor hygiene. In these settlements, only 57% of pregnant women complete the cycle of recommended antenatal care (ANC) visits, and <50% have a safe, assisted delivery; more than half of the women who have complications during pregnancy or peripartum period do not receive medical assistance.^4^

High MMR and NMR have been partly attributed to limited availability of basic ultrasonography and the missed opportunity to tap the diagnostic potential of such technology in monitoring pregnancies.^5^ Ultrasound scans are pain-free, fast and believed to hold no health risk to the mother and fetus.^6^ Diagnostic ultrasonography improves detection of foetal and placental anomalies, both frequent causes of stillbirths.^7,8^ Recent improvements in ultrasound probe technology have significantly reduced costs of machines, and high-quality imaging can be performed using robust, portable devices that can be deployed in challenging, resource-limited settings.^9,10^

Despite the availability of ultrasonography equipment however, a shortage of healthcare providers trained on ultrasonography remains a barrier to access.^11^ A majority of public and private primary healthcare clinics (PHCs) in LMICs lack this essential diagnostic service, and where available, the costs often are prohibitive hence many expectant mothers fail to access these services. While ultrasound services are culturally acceptable and have significant demand in Kenya, economic and geographic obstacles to the use of sonography in pregnancy still persist.^12^ Promoting a wider and more equitable access to obstetric ultrasound scans may make it possible to identify, at early stages, a larger number of conditions that put maternal and newborn survival at risk, and reduce preventable morbidity and mortality.^13,14,15^

Given the shortages of skilled sonographers, and widespread availability of trained nurses and midwives, nurse-led delivery of point-of-care obstetric ultrasound scans has the potential to greatly improve patient care in resource-constrained settings.^17,18^ However both design of nurse training and existing regulations around delivery of ultrasonography services (restricting it to certified sonographers or specialist obstetricians and radiologists) limit the ability for nurses to deliver obstetric ultrasonography services at scale in sub-Saharan Africa.^5,17,18^

Advances in telemedicine and tele-radiology provide an innovative approach to increasing access to scarce, specialized skills in healthcare at considerably lower costs, with the potential of reducing referrals, improving service efficiency, and increasing morale among health workers in low-resource settings.^14,19^

Combining nurse-led, point-of-care ultrasonography with telemedicine review of scan data and images by qualified sonographers could improve the acceptability of nurse-led ultrasonography across diverse stakeholders. This is a key step towards widespread adoption and scaling of nurse-led obstetric ultrasonography across low- and middle-income countries (LMICs), and their drive towards Universal Health Coverage (UHC).^20^

Such a “task shifting” approach however requires multi-stakeholder engagement, training, quality assurance, and deliberate program design to ensure both regulatory compliance and access to high-quality care.^21,22^ In this paper, we explore the acceptability, utility and potential for scale for the nurse-led, point-of-care obstetric ultrasonography services coupled with telemedicine for remote expert reporting.

## Methods

SonoMobile was a three-year project implemented in Nairobi’s urban informal settlements with the aim of improving universal access to antenatal ultrasonography. The formative process evaluation reported here adopted a mixed-methods approach, using both qualitative approaches to draw insights from a range of stakeholders of antenatal ultrasonography, and quantitative methods to analyse programme data repositories.

### Intervention

The intervention to provide ultrasonography services in PHCs offering ANC (**Figure 1**), involved the following components: a) training of nurse midwives via a new curriculum developed in collaboration with the Kenya Medical Training College (KMTC) on nurse-led obstetric ultrasonography; b) antenatal ultrasound imaging at point-of-care by trained nurse midwives at partner PHCs using mobile ultrasound probes; c) remote reporting of ultrasound images by qualified sonographers accredited in Kenya, using telemedicine technology described earlier;^16^ d) quality control of the service by obstetricians through partnership with Italian universities.

**Figure 1:**
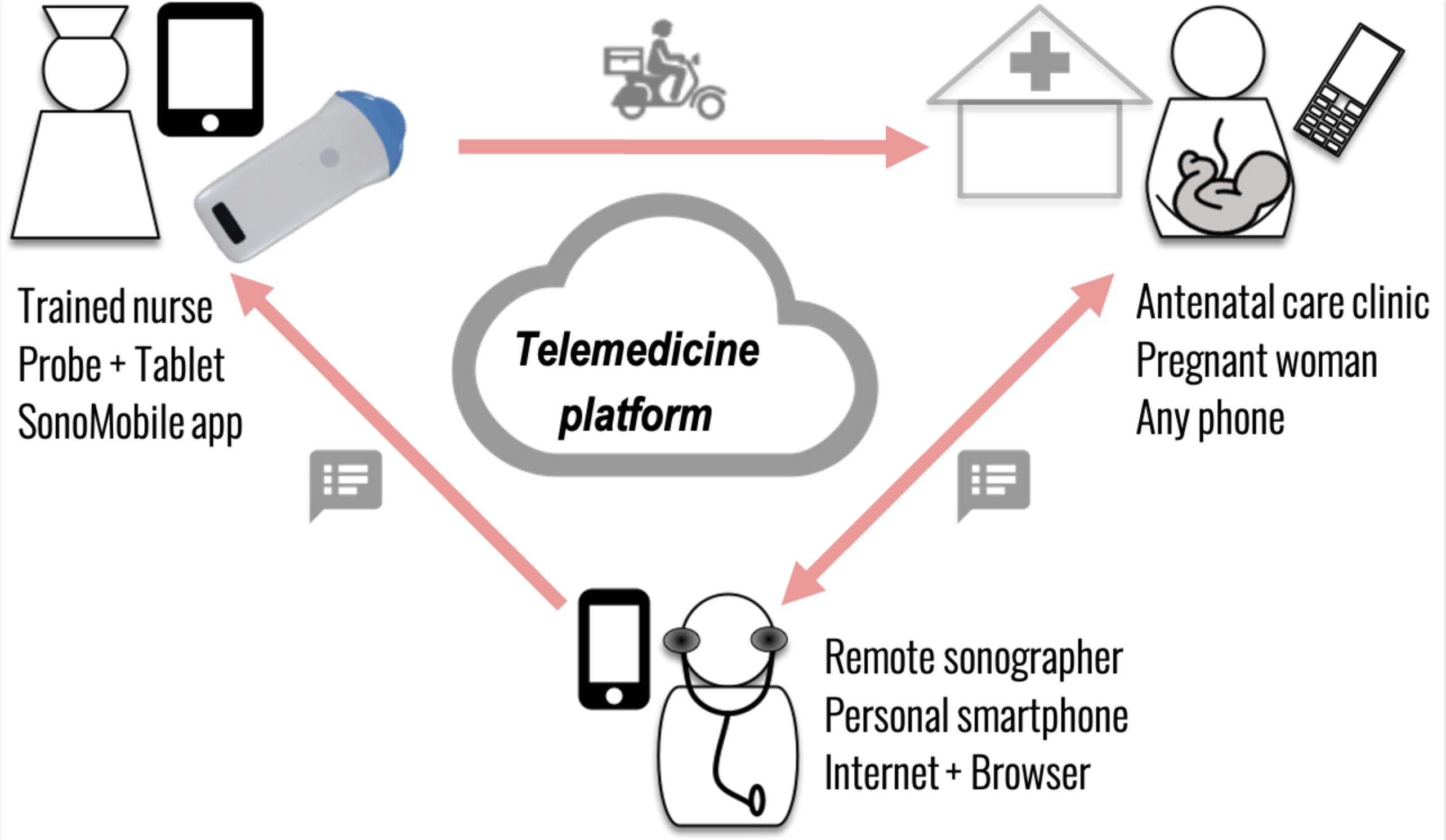
Schematic of Sonomobile intervention.

### Study Site and Sample Selection

The project was implemented in nine Level 2 PHCs in Ruaraka Sub-County in Nairobi County offering ANC services to pregnant women. These PHCs, managed either by the public or private/non-governmental sector, were selected as the main link facilities referring maternity patients to Ruaraka Uhai Neema Hospital (RUNH), run by World Friends Onlus who sponsored the study. These clinics primarily cover catchment populations in the informal settlements in Ruaraka. The provision of point-of-care obstetric ultrasonography services began in October 2019, and the evaluation was conducted in between July 2020 and November 2021.

Purposive sampling was employed in selection of the stakeholders given their respective roles in the implementation, utilization and regulation of the intervention. Insights were drawn from 61 key stakeholders involved in the project (**Table 1)**.

**Table 1:**
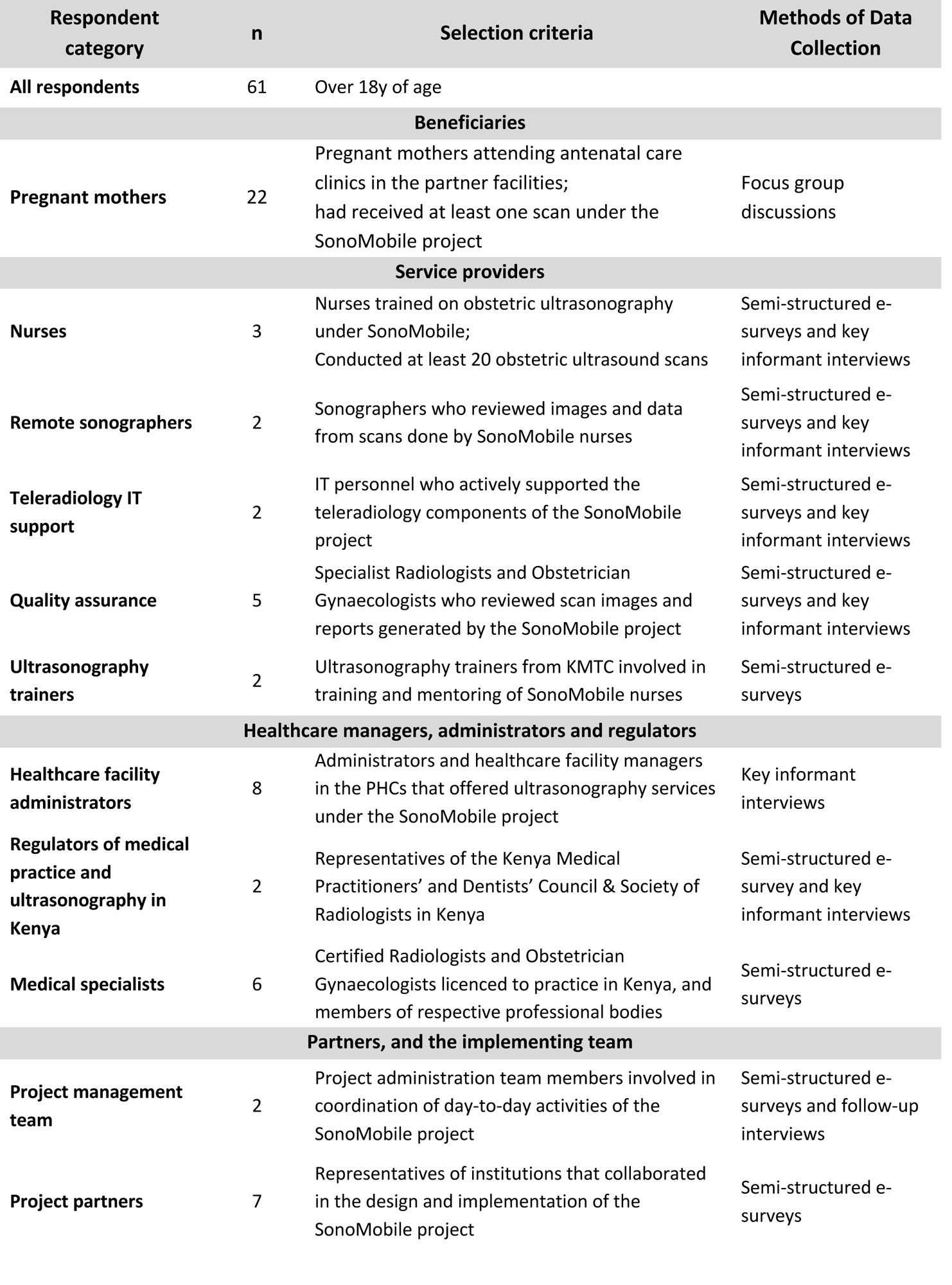
Summary of respondents and data collection methods.

### Data Collection

Qualitative data collection aimed at capturing the experience, suggestions and opinions of the respondents around the following key aspects of the intervention: training, task shifting, telemedicine, service delivery, quality. Qualitative data was collected through key informant interviews (KIIs), semi-structured electronic surveys (e-survey), and focus group discussions (FGDs). KIIs were conducted in person and via phone/internet due to restrictions imposed to contain the Covid-19 pandemic during various periods of the evaluation.

Qualitative data collection tools were developed using conceptual frameworks related to training, task-shifting, telemedicine, scalability and utility; the data collection tools used for each respondent type are also available (**Appendix 1**). The interviews and FGDs, upon obtaining informed consent, were audio recorded via smartphone applications, transcribed, translated as necessary, and stored. E-surveys were shared by email and completed responses were de-identified before analysis.

Quantitative data on the scans conducted and reported were collected by the project implementation team as part of the service delivery, and were de-identified for retrospective review. Scan-level data included provider, site, gestational age, referral, etc. The SonoMobile scanning platform structured each scan by “blocks” which corresponded to a protocol for obstetric ultrasonography. Blocks included, for example, number of foetuses, foetal lie, heart, head and spine, placenta, amniotic fluid, maternal anatomy. Scan images and data were organised by blocks, and block data reported by nurse and sonographer were compared.

Specialist radiologists and obstetricians based in Italy conducted monthly reviews of de-identified scans and sonographer reports. Feedback was provided to the nurses and sonographers in periodic virtual meetings. This study surveyed three remote specialists, but did not access the feedback provided to the nurses and sonographers.

### Data Analysis

First-level analysis and coding of the qualitative data was done through inductive coding (code frame available in **Appendix 2**). Subsequent level analysis was done through deductive and axial coding with establishment of linkages in keeping with the defined frameworks of analysis. The analysis adopted a flat coding frame with the respective codes having similar significance.

Quantitative data were analysed using MS Excel. We report descriptive statistics including the number of ultrasound scans done, scans by gestational age, pregnancy type as well as the number of high-risk pregnancies referred to RUNH for management.

### Ethical Approval

Ethical approval was obtained from the Strathmore University Ethics Review Committee (#SU-IERC0795/20), and a research permit obtained from Kenya’s National Commission for Science, Technology & Innovation (#827569).

## Results

Six nurses were trained on basic concepts in obstetric ultrasound imaging at RUNH between Jun 2019 and Jul 2020; the curriculum developed by the SonoMobile project in collaboration with the KMTC was later approved is currently delivered as a 6-month short course on obstetric ultrasonography for nurses at KMTC Nairobi. A total of 4,567 point-of-care obstetric ultrasound scans were conducted under the SonoMobile project between Oct 2019 and Aug 2021 in nine PHCs in the informal settlements of Ruaraka, Nairobi County, Kenya. Scan images and data were reviewed by sonographers employed by RUNH. As context to the results presented here, **Figure 2** illustrates, where data were available, the number of new ANC visits, the proportion of mothers completing at least four ANC visits, and the proportion of ANC visits occurring in the first of pregnancy in Ruaraka sub-county where SonoMobile was implemented.

**Figure 2:**
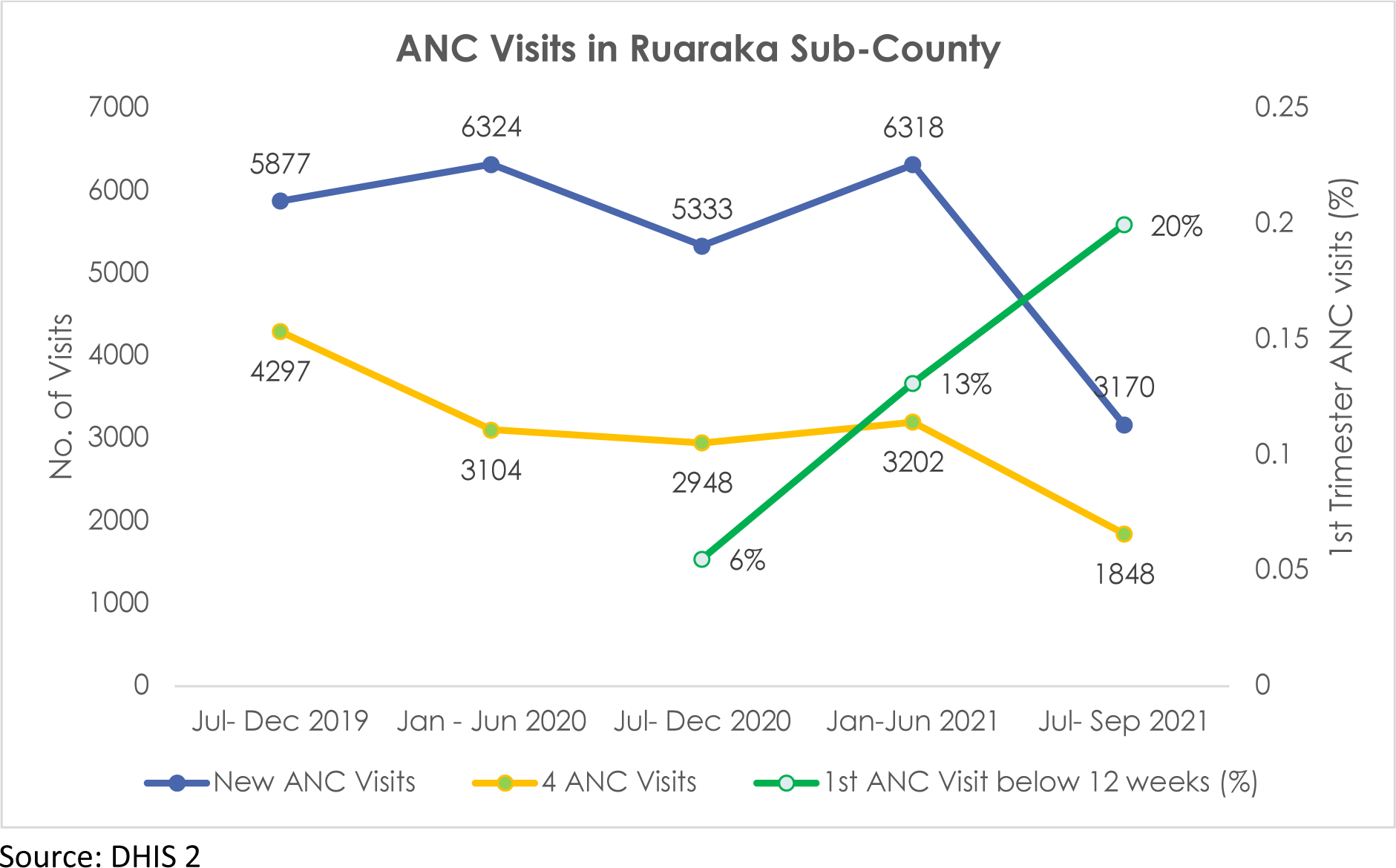
ANC Utilization in Ruaraka Sub-County.

### Value of the task-shifting service

All respondents sampled across all respondent categories saw value in the intervention and expressed interest in using and expanding it. The value of the service was described in the following main categories: increasing geographic and financial access to services; improving technical and experiential quality of ANC; improving awareness of value of antenatal ultrasound and more generally, ANC among pregnant mothers; and informed decision-making for high-risk pregnancies. These are described in further detail with representative quotes in **Table 2**.

**Table 2:**

| Perceived value of intervention | Respondent categories voicing this value | Representative quote(s) | Contradicting minority opinions |
| --- | --- | --- | --- |
| Improved geographical access to antenatal ultrasound services | All categories | ... in such areas like Mathare, Baba Dogo, Redeemed, they don't have the ultrasound machines. But now this project came up with those machines where they can access them.<br>- **Sonographer 1** |  |
| Improved financial access to antenatal ultrasound services | Mothers<br>Health workers<br>Project administrators<br>Regulators | "It has been a very important program actually to our villagers who most of them are poor. So, they have actually benefited from this because if you go all over, the ultrasound scan is expensive."<br>- **Health Worker/ Administrator (Partner Facility 2)**<br>"We are surprised and shocked by the number of women who were willing to pay for it as a service."<br>- **Project Administrator** | Affordability after introduction of the service charge (500 – 1000 KES) was still a concern and impacted demand. Integration with existing financial access programmes (eg. Linda Mama, NHIF) is an opportunity |
| Increased awareness of importance of antenatal ultrasound | Mothers<br>Health workers<br>Project nurses | There are improvements because we now see mothers have knowledge about the scan. At least now the mothers are getting aware. Because they teach one another.<br>- **Health Worker, Partner Facility 2**<br>Once they heard of it...the mothers that were attended to and had an experience with that (the project) and knew that it was important for them ....., they would go out in the community and spread the news. And other mothers would come telling us that, 'we heard this project from our friends who had experienced it.'"<br>- **Project Nurse 2** |  |
|  |  | It helped me to know how the baby is progressing and developing in the womb. If there is any complication you are told. And also knowing the gender of the baby.<br>**- Mother, FGD 2** |  |
| **Alignment with technical quality standards for antenatal care** | Specialist<br>Remote sonographers<br>Regulators | Individuating first level and second level professionals that perform same exams with different insight levels can widen the number of patients offered the ob-scans but narrow the indications to the expert sonographer referral, ultimately leading to a more precise and better quality of care.<br>**- Specialist 1, QA Team**<br>It does fit because these are the same people who assist women in delivery. They are the same people that take these mothers throughout their pregnancy journey. So, them getting a skill where they are not only able to touch but also use something that helps them to see...I feel it is sort of balanced. This is someone who is aware of what is going on in terms of pregnancy and the baby and then someone who is skilled in reporting on the images gives their own feedback on the same.- **Project Administrator 2** | The regulators and trainers sampled reiterated that the point of care scans were neither a replacement for nor synonymous to the full abdomen and pelvis scans as the latter were beyond the scope of the nurses' training. |
| **Improved patient experience aspects of quality of antenatal care – patient centredness** | Mothers<br>Health workers<br>Facility administration<br>Project administration | We were treated well, and they talked to us well.<br>**- Mother, FGD 2**<br>In this case they were the ones going to the patients. Once you are exposed to that, you now understand what a patient goes through past what they can tell you in a hospital set up. It's more personal and interactive.<br>**- Project Administration** | A few respondents highlighted the challenges that reduce the patient-centeredness of the services including service delays, lack of user-friendly outputs/ reports, limited availability at facilities, and rejection of the |
|  |  | You know they know that our scan is on Wednesday, so you find that they come within the week and are booked for Wednesday.<br>**- Facility Administrator, Partner Facility 2**<br>From the experience I have with SonoMobile, the prices are friendly and the mothers have an interactive session with the sonographer to explain to them the results.<br>**- Health Worker, Partner facility 4**<br>There is a day I received a message. But the others have never called me to go to scanning or send me a message. But for this one, there was a message.<br>**- Mother, FGD 3** | results and reports by other facilities while questioning their legitimacy. |
| Improved capacity for nurses within the scope of their scheme of service | Health workers<br>Ultrasonography trainers<br>Regulators | It is not a new cadre of staff since it is just point of care ultrasonography. As such, existing laws and under the respective Acts (Health Act 2017, Medical Practitioners and Dentists Board Act CAP 253, Nurses Act CAP 257) and related guidelines by the Medical Council, Nursing Council are sufficient.<br>**- Regulator 2**<br>No regulatory conflict if this remains as Point of Care. However, if the scope is to be broadened, there may be need for clarity.<br>**- Trainer 2** | While the training of nurses in the current form improved their capacity (knowledge and skill) in point of care obstetric ultrasonography, most health workers, the trainers and the regulators sampled considered it inadequate for comprehensive obstetric ultrasonography due to its limited scope, and a lack of clarity on the regulatory standards for the nurses conducting the scans. |

The majority of the respondents highlighted the improved capacity for health workers, specifically nurse midwives, to assess and manage pregnant mothers appropriately and comprehensively. Where necessary, they would refer pregnant women for specialised care. The value of the training was further affirmed by the trainees (nurses) and their trainers noting its appropriateness in meeting an existing need for obstetric ultrasound in the informal settlements of Ruaraka.

Respondents in facilities and pregnant mothers were particularly sensitive to the cost of the services against its perceived value. While most scans were provided free of cost, a scan fee of between five hundred to one thousand shillings (∼$4-$8) was introduced in Oct 2021 by some private sector PHCs, which was also the indicative cost provided in the FGDs. Many noted that the service was relatively affordable for the target population in the informal settlements, with most mothers willing to pay for the service. However, facilities did see a decline in the rate of mothers getting the scans upon introduction of a service charge (observations from the project management team; no data available). Some concerns persisted in mothers about the effect of ultrasound on the foetus or the mother, and this also affected perceptions of the risk-benefit trade-off of getting a scan.

### Access to, and quality of the service provided

Figure 3a illustrates the time series of the scans performed under the SonoMobile project between Oct 2019 and Aug 2021; an average of 13.4 scans were performed on each day that the service was delivered in that period. The daily average increased from 6.9 in the pre-covid period to 14.6 in the period between July 2020 and Aug 2021 as both the number of PHCs offering antenatal ultrasonography and the demand in PHCs for point-of-care services increased; scan numbers remained steady after that. The utilization of the point-of-care obstetric ultrasound scans (Figure 3b) was lowest during the first trimester of pregnancy (2.6% of all scans conducted in the project), and the majority (>60%) were done in the third trimester.

**Figure 3:**
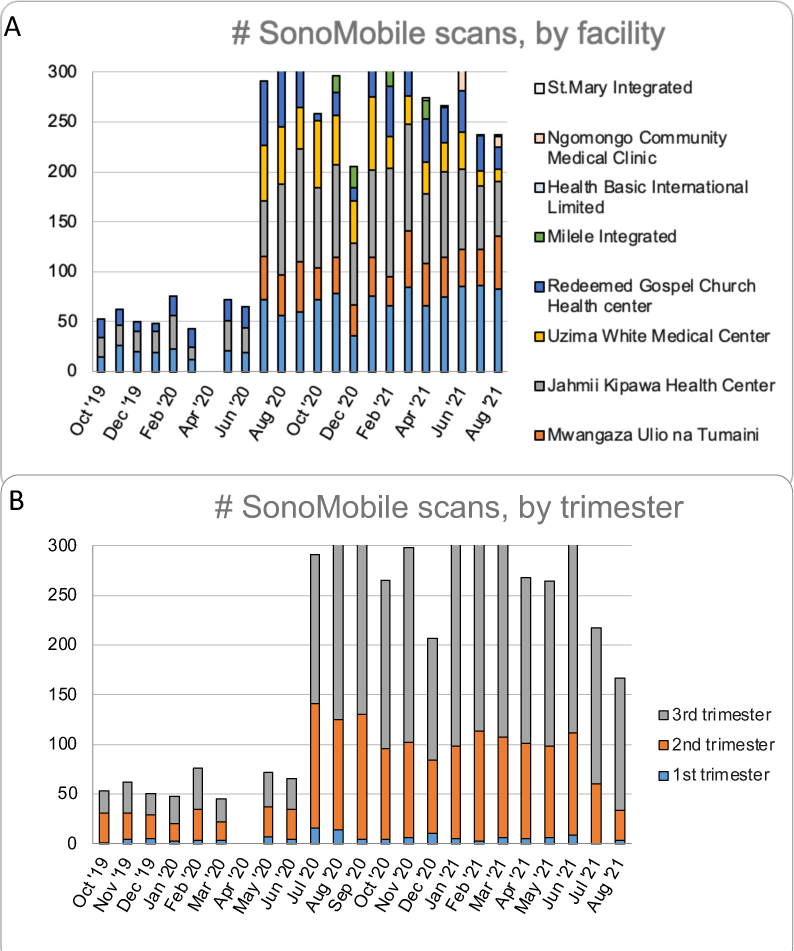
Monthly SonoMobile scans, by facility and by trimester.

The low uptake of first trimester scans is partly explained by limited awareness amongst the pregnant mothers and health workers on the value early obstetric ultrasound scans for both mother and child. A few respondents among pregnant mothers and health workers pointed out the fact that access was limited by services not being available daily in every PHC. Other factors cited included high cost of scans (after scan fee was introduced), distance to facilities with scans, fears and misconceptions on the effect of the scans on the mother and her pregnancy, and fear of wrong reports on gender of the foetus.

### Potential for implementation at scale: Regulation and sustainability

Some of the respondents noted the potential of the service in improving system-wide efficiency by ensuring that only necessary cases were referred to higher level. The value of reduction of referrals from PHCs were highlighted by both mothers and facility administrators. Of the 4,567 scans performed, 779 (17%) resulted in referral for further management. The bulk of the referrals were for multiple pregnancies (33%), followed by placental anomalies (18%). No pregnancy was reason for referral in 18% while no reason was provided in 17%, followed by foetal death and foetal anomalies (10%).

Overall, there was limited awareness of the existence of any regulations and policies that guide the practice of nurse-led ultrasonography, as highlighted by a facility administrator: “ultrasound services are not a nursing procedure so there are no laws governing this.” Even representatives of regulatory bodies were unsure of who should regulate nurse-led scanning and how telemedicine reporting by sonographers were regulated. Task shifting to nurses to conduct scans was opposed by specialists who raised concerns about the scope of training, systems to evaluate of competency, and technological barriers to telemedicine-based reporting.

The following were cited as essential enablers for scaling nurse-led point-of-care obstetric ultrasonography: continuous capacity development of nurse midwives and sonographers; stakeholder engagement in capacity development, service delivery design and implementation; strategic collaborations with other maternal and neonatal health service providers and financing structures; robust quality assurance and supportive supervision; progressive improvement of technologies (hardware and software) used in the service; strong referral systems; clear regulations on training and quality assurance of nurse-led ultrasonography with remote sonographer review.

## Discussion

This study presents findings on acceptability, utility and sustainability of nurse-led, point-of-care obstetric ultrasonography with telemedicine review gathered from an intervention conducted in the informal settlements in Nairobi. It is estimated that 97.6% of pregnant women within Ruaraka sub-county have at least one prenatal visit,^23^ so access to a health facility is not inherently problematic; however, it is estimated that only 27% of pregnant women in this area receive an antenatal ultrasound scan during pregnancy (unpublished data from study sponsor). In this context it is not surprising that all stakeholders of the intervention perceived the value nurse-led point-of-care obstetric ultrasonography in bridging existing barriers to accessing high-quality ANC services within urban informal settlements.

The findings highlight the poor uptake of ANC services, including obstetric ultrasonography, in the first trimester of pregnancy among the population served. Early ultrasound can not only improve early detection of multiple pregnancies and foetal anomalies,^24^ but can also improve gestational dating,^7^ which may result in fewer inductions of labour – a likely contributor to poor maternal and neonatal outcomes.^25^ Future implementations of obstetric ultrasonography in LMICs would ideally focus on increasing rates of ultrasonography in the first trimester.

Overall, there was affirmation and positive feedback from both trainees and trainers on their experience with increasing nurse-led, point-of-care obstetric ultrasonography. While the training of nurse midwives on obstetric ultrasonography was highly rated by the trainees, it was noted to be inadequate for comprehensive obstetric and pelvic imaging.^26^ Trainees also highlighted the need to improve training for first trimester imaging. Scaling nurse-led obstetric ultrasonography is likely to involve a balance between the scope of the training curriculum and training duration. Longer training programmes are likely to be more challenging for both experienced nurse midwives and their employees. Future efforts would balance the scope of training and its duration and explore the role of artificial intelligence (AI) in improving quality of nurse-led ultrasonography,^27^ both during training and through feedback, either in real time or asynchronously.^28^

Respondents broadly felt that the nurse-led, point-of-care obstetric ultrasonography was useful in improving access to an important diagnostic tool in obstetrics, and in reducing the cost of ultrasonography – a broadly accepted view from other studies of point-of-care ultrasonography in LMICs.^18,29,30^ This study expands the understanding of such interventions to include remote review by sonographers using telemedicine. Such remote review is likely to be a key part of future interventions in resource-constrained settings for a number of reasons: a) as discussed above, training of nurse midwives on obstetric ultrasonography is likely to be shorter than current training of clinical officer sonographers; b) shorter training duration is likely to limit the ability of nurse midwives to provide obstetric ultrasonography services at scale; c) quality assurance and accreditation of nurse-led obstetric ultrasonography is likely to need remote, independent review of nurse scans; d) remote review using asynchronous telemedicine models are likely to be more appropriate and easier to implement in low-resource settings with limited internet connectivity and bandwidths and culture of technology adoption than standalone, AI-based models for quality improvement.^19,27^

Combining nurse-led point-of-care ultrasonography with remote reporting of scans by clinical officer sonographers describes a task-shifting exercise, which is well understood in LMIC settings.^22^ Here, it shifts the responsibility of scanning to nurses, the most numerous cadre in LIMC health systems and responsible for almost all skilled deliveries.^31,32^ The efforts of clinical officer sonographers (and possibly of specialist radiologists and obstetricians) are focused on reviewing and reporting scan images and data, identifying high-risk pregnancies, and supporting nurses in patient management and referral. This separation of roles using telemedicine is a well understood strategy to overcome shortages in human resources for health in LMICs.^19,31,33,34^ While well-understood at small scale, task-shifting of obstetric ultrasonography to nurse midwives requires new and continued investments in technologies and health worker capacity for effective provider-provider interactions,^19^ and linkage to existing financing mechanisms aligned to UHC.^35^

### Generalisability and Limitations

The intervention and the study were affected significantly by the pandemic. While the intervention itself was forced to stop during lockdowns, the value of point-of-care ultrasonography at PHCs close to pregnant mothers was highlighted by the pandemic. However, scheduling and conduct of participant interviews and focus groups were challenging.

Like with many complex interventions, it is difficult to assess the impact of nurse-led obstetric ultrasonography on downstream outcomes like maternal and neonatal mortality and morbidity.^36^ It was beyond the scope of this study to measure the outcome of targeted referrals, skilled delivery or timing of first antenatal care visits – all likely process-level changes resulting from increased access to point-of-care ultrasonography.

### Recommendations and Conclusions

Nurse-led point-of-care ultrasonography is being increasingly utilised across LMICs with various levels of training, support and supervision. The use of telemedicine to enable sonographer review of images and data captured by nurses can enable effective regulation of the service – a key step towards scaling and sustainability. The role of telemedicine in the context of task shifting is well understood, and this technology is likely to play a central role in delivering universal access to a life-saving technology across LMICs.

## Data Availability

All data produced in the present study are available upon reasonable request to the authors

## Acknowledgements

The authors would like to thank Jacopo Rovarini for his contribution to intervention design and fundraising.

## Author contributions

CMM led on developing tools, collecting data and first round of analysis and drafting of thematic findings. MBK led further analysis and drafting of full paper. GFM and PK secured funding for the study. PK led on intervention design and strategic directions for data collection and analysis. GFM, MVD, LP, SS and YG supported data collection and access to field sites. All authors reviewed the draft paper and agreed on the submitted version.

